# Local Control Comparison of Early-Stage Epidermal Skin Cancers Treated With and Without Dermal Image Guidance: A Meta-Analysis

**DOI:** 10.1101/2022.09.19.22280122

**Authors:** Lio Yu, Mairead Moloney, Alison Tran, Songzhu Zheng, James Rogers

## Abstract

**Background:** Various treatments exist for non-melanoma skin cancer (NMSC), but the mainstay is surgical removal. Superficial radiotherapy (SRT) is one non-surgical technique that has been used for over a century but fell out of favor due to the advent of Mohs micrographic surgery (MMS). A new technology that combines a 22 megahertz (MHz) dermal ultrasound with SRT (US-SRT) enables tumor visualization before, during, and after treatment, and demonstrates increased cure rates and reduced recurrences.

**Methods:** We conducted a meta-analysis comparing the local control (LC) of four studies using traditional non-image-guided forms of radiotherapy for NMSC treatment to two seminal studies utilizing high-resolution dermal ultrasound-guided SRT (HRUS-SRT). The four traditional radiotherapy studies were obtained from a comprehensive literature search used in an article published by the American Society of Radiation Oncology (ASTRO) on curative radiation treatment of basal cell carcinoma (BCC), squamous cell carcinoma (SCC) and squamous cell carcinoma in-situ (SCCIS) lesions. The meta-analysis employed a logit as the effect size indicator with Q-statistic to test the null hypothesis.

**Results:** LC rates for the 2 US-SRT studies were statistically superior to the 4 traditional therapies individually and collectively. When stratified by histology, statistically superior outcomes for US-SRT were observed in all subtypes with p-values ranging from p < 0.0001 to p = 0.0438. These results validated an earlier analysis using a logistic regression statistical method showing the same results.

**Conclusion:** US-SRT is statistically superior to non-image-guided radiotherapies for NMSC treatment. This modality may represent the future standard of non-surgical treatment for early-stage NMSC.

## 1.0. Introduction

Non-melanoma skin cancer (NMSC) is the most prevalent cancer in the United States (US) [1]. Despite its prevalence, NMSC is generally not reported or tracked by national cancer registries [2]. The most current data from 2012, estimated that there were 5.43 million NMSC lesions amongst 3.32 million individuals, with a substantial portion of people having more than one lesion [3].

Epidermal NMSC comprise the overwhelming majority (99%) of lesions with basal cell carcinoma (BCC) contributing to 80% of cases and squamous cell carcinoma (SCC)/squamous cell carcinoma in-situ (SCCIS) comprising the remaining 20% [4]. NMSC lesions typically occur in individuals 60 years or older and are found on sun-exposed areas of the body [4, 5]. Although mortality and metastasis from these cancers are uncommon, treatment remains the standard of care [2, 6].

NMSC can be treated in numerous ways, both surgically and non-surgically. The “gold standard” treatment in the US is Mohs micrographic surgery (MMS) [1]. Radiotherapy (RT) is a non-surgical modality that has been used for over a century for the treatment of skin cancer, but has fallen out of favor with the advent of MMS [7, 8]. However, radiotherapy provides advantages in cosmetic and functional outcomes to patients as NMSC lesions predominantly affect cosmetically sensitive areas, such as the face, nose, and ears [8]. Furthermore, recent advancements incorporating imaging into radiotherapy result in excellent local control (LC) rates similar to MMS [8, 10, 16].

Recently there has been a resurgence in the use of RT, specifically, superficial radiotherapy (SRT) in dermatology offices. Within the last decade, the addition of high-resolution dermal ultrasound (HRUS) to SRT units has enhanced NMSC treatment by adding a high-frequency (22 megahertz) dermal ultrasound probe directly to the SRT base unit to visualize the layers of the skin. HRUS allows clear visualization differentiating between the epidermis, dermis, and subcutaneous fat thereby facilitating the determination of NMSC tumor size in all axes to visualize the NMSC lesions prior to treatment [9]. This ultrasound (US) technology can also be used during and after treatment. This dermal US capability has led to the development of ultrasound-guided superficial radiation therapy (US-SRT). US-SRT started use in approximately 2015 and is also known in other studies as image-guided superficial radiotherapy (IGSRT) [10, 16].

This report compares this newer novel technology, US-SRT, to traditional non-image-guided radiotherapy treatments of NMSC, namely, external beam radiation therapy (XRT) and SRT. A recent logistic regression approach examining the same datasets in this paper was recently submitted for consideration for publication. Since the methodology is entirely different than a meta-analysis approach, this paper serves to verify or refute the findings of the logistic regression article.

### 1.1 Objective

To statistically compare the recurrence rate between HRUS image-guided SRT versus non-image-guided radiotherapy modalities for the treatment of early-stage NMSC.

## 2.0 Methods

### 2.1 Study Design: Traditional Radiotherapy Study Selection

The American Society of Radiation Oncology (ASTRO) published a literature review on curative radiation treatment of BCC and SCC lesions and reviewed 143 studies over 30 years, from 1988 to 2018. Figure 1 outlines the specific inclusion and exclusion criteria used to select comparable, large sample (*n* = >100), American studies that utilized SRT and/or XRT. Only four recent, evidence-based and pertinent for comparison to US-SRT studies met the inclusion/exclusion criteria. These studies are Lovett, et al. 1990, Locke et al. 2001, Silverman et al. 1991, and Cognetta et al. 2012 [11, 12, 13, 14].

**Figure 1.**
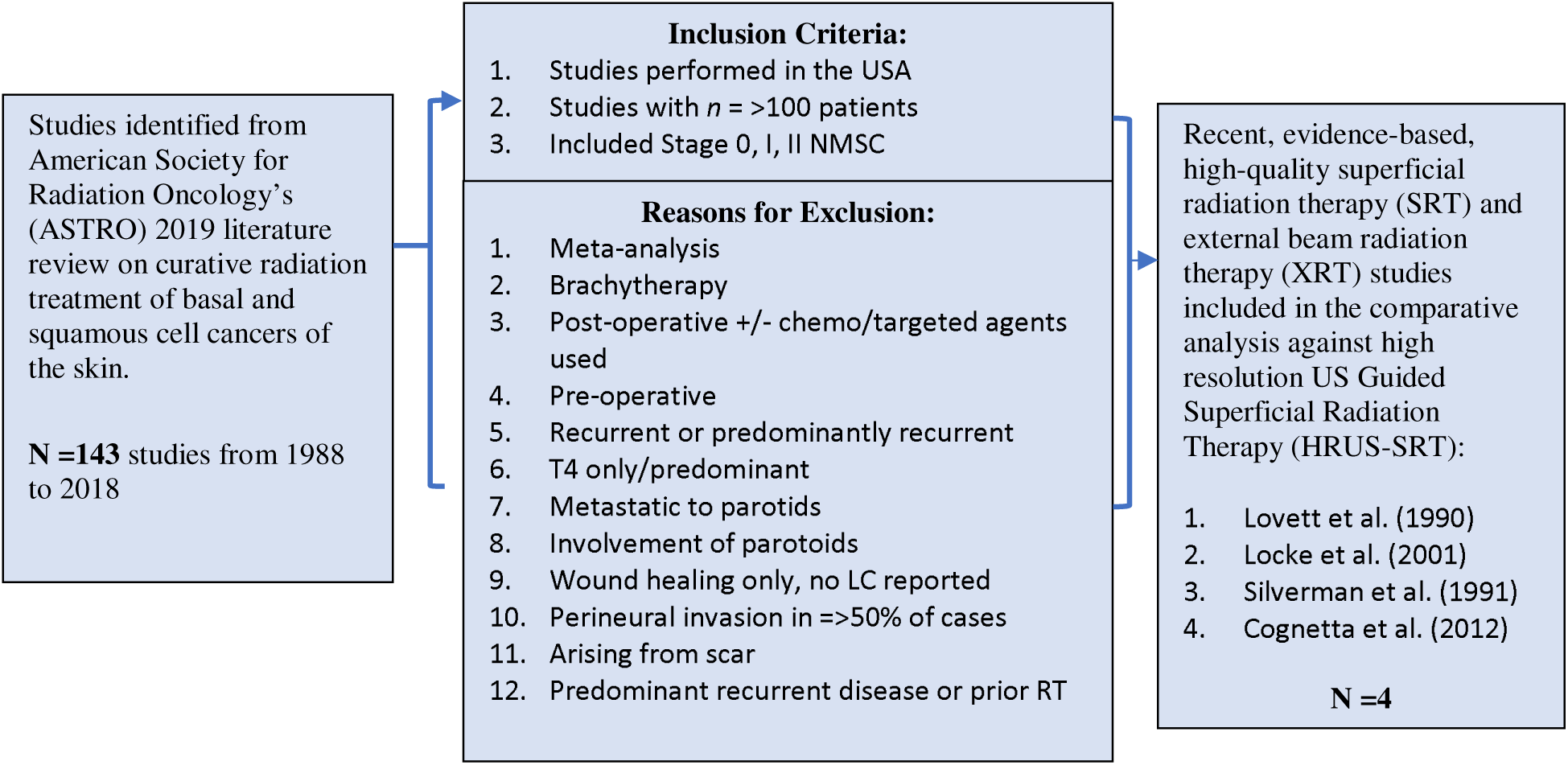
Flow chart of the selection process and reasons for exclusion resulting in identifying four studies from the American Society of Radiation Oncology (ASTRO) published literature review on curative radiation treatment of BCC and SCC lesions.

The main objective was to select large sample studies having NMSC lesions with comparable histology and stages (i.e., stage T_is_, T_1_, T_2_ lesions) to compare “apples to apples.” This resulted in reasons for exclusion listed in Figure 1.

### 2.2 Novel Technology, US-SRT, Study Selection

We searched PubMed between inception and July 1, 2022, for studies (key search terms: “((image-guided superficial radiation therapy) AND (skin cancer)”) and searched other sources of public information, such as Google Scholar. We identified only two published studies that utilized US-SRT to treat NMSC, and both studies include one of our co-authors – Dr. Lio Yu. The large 2021 study by Yu et al. treated 2917 histopathological confirmed NMSC lesions in 1632 patients with US-SRT [10]. This study was utilized in this analysis and will be referred to as the Yu 2021 US-SRT study. The other published study utilizing US-SRT treated 133 NMSC lesions in 94 patients [15]. A recently nationally presented study that combined data from the two published papers as well as updated follow-up (mean follow-up of 2.1 years and maximum follow-up of 6.5 years) was identified and published only in abstract form. This abstract, referred to here as the Moloney 2022 US-SRT study [16], included 1725 patients with 3050 lesions and was utilized as one of the 2 studies used in the US-SRT arms in the current meta-analysis.

The Yu 2021 US-SRT study included data collected from seven outpatient dermatology clinics across the United States. The Moloney 2022 US-SRT study utilized the same data with additional data from one other outpatient dermatology practice and methodology as the Yu 2021 US-SRT study [16]. The study design consisted of a retrospective chart review of patients with histopathological confirmed NMSC lesions treated with US-SRT at each clinic. Treatment data and lesion characteristics were collected and documented. Lesions included in both studies were stage 0, I, or II (i.e., Tis, T1, or T2 without clinical evidence of regional lymph node or distant disease (N0 and M0)) at presentation, based on the American Joint Committee on Cancer (AJCC) 8th Ed. Cancer Staging Manual [17]. The AJCC 8th Ed. staging pertained to cutaneous SCC of the head and neck; these same criteria for staging were applied in the Yu and Moloney studies to all BCC, SCC, and SCCIS lesions throughout the entire body for consistency. Lesion data, treatment data, and follow-up intervals were initially gathered manually from written and electronic medical records. Updates to patient follow-up intervals were accessed electronically with the assistance of algorithmic analysis provided by a healthcare data company (Sympto Health, Inc.). The local control results from the four studies identified in the ASTRO literature review that utilized traditional non-image-guided radiotherapy were compared to the two US-SRT studies.

The authors of the Yu 2021 and Moloney 2022 studies adhered to the principles established in the Federal Policy for the Protection of Human Subjects, referred to as the “Common Rule,” as well as the pertinent sections of the Helsinki Declaration and its amendments. The data was de-identified for use in their studies.

### 2.3 Meta-analytical Methodology

The four comparator studies (Locke, Lovett, Silverman, and Cognetta) evaluated different subject samples and were therefore independent of each other, save for Locke and Lovett, which were from the same institution (Mallinckrodt), but published 11 years apart and may have had overlapping patients (not explicitly stated in the papers and were treated as independent studies), whereas, the Yu and Moloney studies contained overlapping subject samples, and thus, exhibited correlated outcomes. To adhere to the meta-analytical requirement of independence among studies, two separate meta-analyses were necessary, one using the Yu sample and one using the Moloney augmented sample to assess US-SRT relative to XRT/SRT used in the comparator studies. Note that the augmentations inherent in the Moloney sample have been described in a previous section and would be expected to explain differences, if any exist, between the two meta-analyses (Yu v. Moloney as the US-SRT comparator). Each meta-analysis employed the logit (i.e., the natural log of the odds) as the effect size indicator. For convenience, the corresponding local control event rate and local control odds of event have been added to the statistical presentation. Thus, the outcome for each US-SRT and each XRT/SRT study sample may be conceptualized as an event rate, an event odds, or an effect size logit. A forest plot presenting the logit effect sizes with 95% confidence intervals by type of lesion for each US-SRT study and each XRT/SRT study was constructed. A Q-statistic was used to test the null hypothesis of homogeneity of logit outcome for US-SRT v. XRT/SRT comparisons with rejection of the null (i.e., p < 0.05) indicating a statistically significant difference between US-SRT and XRT/SRT. Comparisons were carried out for all lesions (BCC, SCC, and SCCIS) combined as well as for each cancer type separately. The statistical comparisons consisted of each US-SRT study compared to all XRT/SRT comparator evaluations with all lesions combined (Figure 2); and of each XRT/SRT comparator compared to both the combined and separate US-SRT studies by lesion type (Table 1).

**Figure 2.**
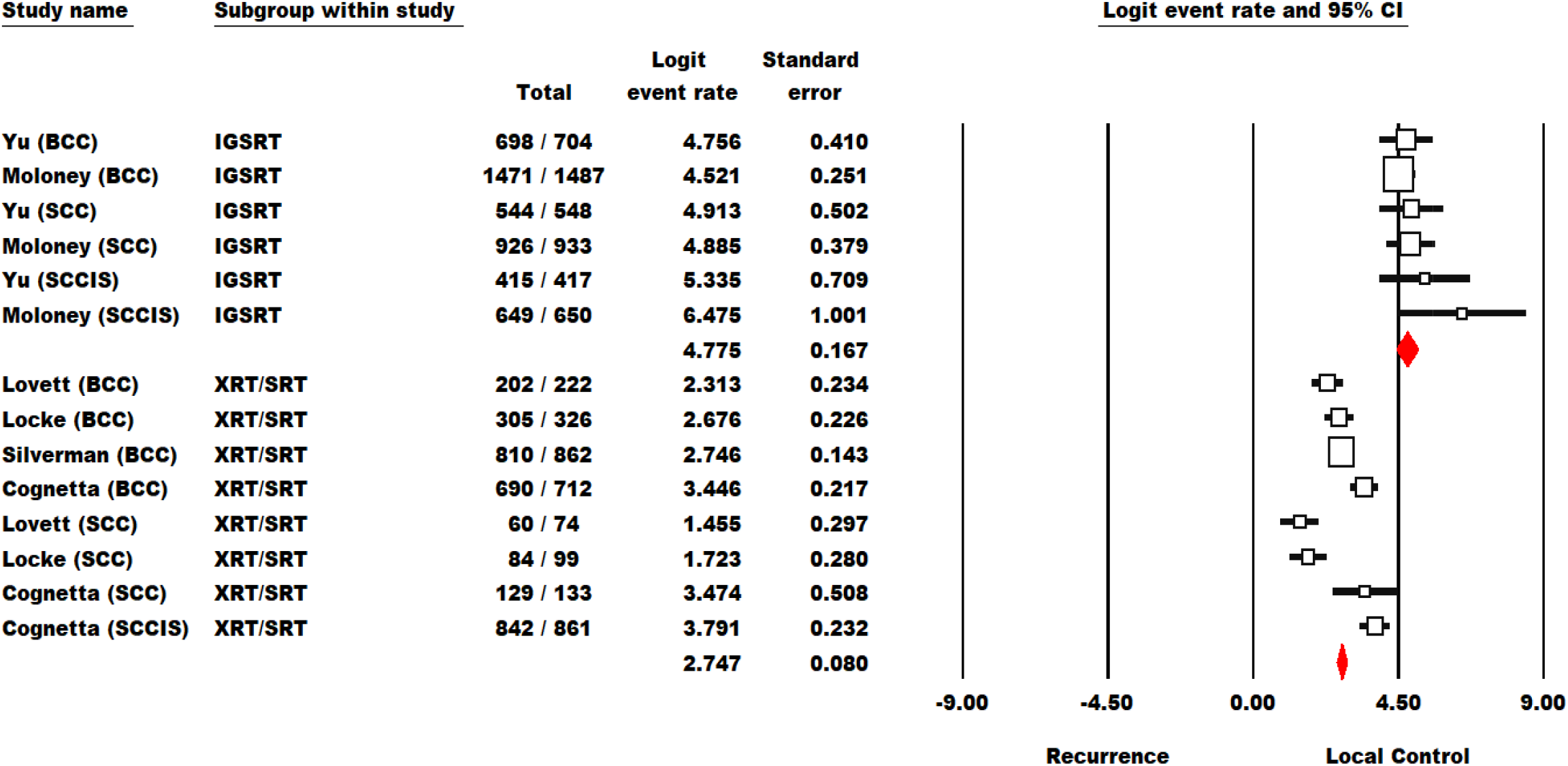
Overall Meta-analytical Comparison of US-SRT and XRT/SRT Outcomes in the Treatment of Basal Cell Cancer (BCC), Squamous Cell Cancer (SCC) and Squamous Cell Cancer in Situ (SCCIS) **<u>All Lesions Contrasts:</u>** **Yu US-SRT v. XRT/SRT Studies Combined, Q [1] = 51.5, p < 0.0001** **Moloney US-SRT vs. XRT/SRT Studies Combined, Q [1] = 79.3, p < 0.001** Note: The displayed overall effect size for US-SRT contains correlated outcomes (i.e., shared patients in part across investigators). The common effect size for XRT/SRT outcomes contains independent outcomes with the rare exception of a few patients who present with more than one carcinoma type. Therefore, separate analyses are presented for the US-SRT investigators, Yu and Moloney, to maintain the assumption of within-analysis independence.

**Table 1.** Outcomes and Meta-analytic Contrasts (US-SRT v. XRT/SRT) for Basal Cell Cancer (BCC), Squamous Cell Cancer (SCC) and Squamous Cell Cancer in Situ (SCCIS)

| Lesion | Treatment | Study | Outcome |  |  |  |  |  | Q Test for Heterogeneity<br>(Contrast: US-SRT v. XRT/SRT) |  |  |  |
| --- | --- | --- | --- | --- | --- | --- | --- | --- | --- | --- | --- | --- |
|  |  |  | Events<br>(N) | Recurrence<br>(n) (%) |  | Event<br>Rate<br>(%) | Event<br>Odds | Effect<br>Size<br>(Logit<br>) | Yu |  | Moloney |  |
|  |  |  |  |  |  |  |  |  | Q | p-value | Q | p-value |
| BCC | US-SRT | Yu | 704 | 6 | 0.9 | 99.1 | 116.7 | 4.76 |  |  |  |  |
|  |  | Moloney | 1487 | 16 | 1.1 | 98.9 | 91.8 | 4.52 |  |  |  |  |
|  | XRT/SRT | Lovett | 222 | 20 | 9.0 | 91 | 10.1 | 2.31 | 26.8 | <0.0001 | 41.3 | <0.0001 |
|  |  | Locke | 326 | 21 | 6.4 | 93.6 | 14.6 | 2.68 | 19.8 | <0.0001 | 29.8 | <0.0001 |
|  |  | Silverman | 862 | 52 | 6.0 | 94 | 15.6 | 2.75 | 21.4 | <0.0001 | 37.7 | <0.0001 |
|  |  | Cognetta | 712 | 22 | 3.1 | 96.9 | 31.5 | 3.45 | 8.00 | 0.0047 | 10.5 | 0.0012 |
|  |  | All | 2122 | 115 | 5.4 | 94.6 | 16.4 | 2.80 | 21.6 | <0.0001 | 41.0 | <0.0001 |
| SCC | US-SRT | Yu | 548 | 4 | 0.7 | 99.3 | 135.6 | 4.91 |  |  |  |  |
|  |  | Moloney | 933 | 7 | 0.8 | 99.2 | 131.6 | 4.88 |  |  |  |  |
|  | XRT/SRT | Lovett | 74 | 14 | 18.9 | 81.1 | 4.3 | 1.46 | 35.2 | <0.0001 | 50.7 | <0.0001 |
|  |  | Locke | 99 | 15 | 15.2 | 84.8 | 5.6 | 1.72 | 30.8 | <0.0001 | 44.9 | <0.0001 |
|  |  | Cognetta | 133 | 4 | 3.0 | 97 | 32.1 | 3.47 | 4.06 | 0.0438 | 5.0 | 0.0260 |
|  |  | All | 306 | 33 | 10.8 | 89.2 | 6.4 | 1.86 | 32.5 | <0.0001 | 51.0 | <0.0001 |
| SCCIS | US-SRT | Yu | 417 | 2 | 0.5 | 99.5 | 208.5 | 5.34 |  |  |  |  |
|  |  | Moloney | 650 | 1 | 0.2 | 99.8 | 652.0 | 6.48 |  |  |  |  |
|  | XRT/SRT | Cognetta | 861 | 19 | 2.2 | 97.8 | 44.3 | 3.79 | 4.3 | 0.0385 | 6.83 | 0.0090 |

Notes: LC = Local Control. Degrees of Freedom = 1 for all US-SRT v. XRT/SRT contrasts. Logits were converted to odds by exponentiation. Due to overlapping study populations, contrasts were conducted separately for Yu and Moloney to maintain the assumption of within-analysis independent outcomes.

## 3.0 Results

Figure 2 presents a forest plot that contains the US-SRT logit effect sizes with a 95% confidence interval by lesion type for each US-SRT study and each XRT/SRT study. By observation, the US-SRT logits are generally superior to the XRT/SRT logits. When lesions are combined and compared between each US-SRT study (i.e., the Yu study and the Moloney study) and all XRT/SRT studies combined, US-SRT is found to be statistically superior to XRT/SRT, and evidence a superior outcome to the combined XRT/SRT (Yu, p < 0.0001 and Moloney, p < 0.0001).

Table 1 compares, by lesion type, US-SRT for Yu and Moloney to the XRT/SRT studies separately and combined. Note that not all XRT/SRT studies evaluated all cancer types and that Table 1 contains, for a given cancer only, those XRT/SRT studies that considered that cancer. In all instances, a statistically superior outcome for US-SRT compared to XRT/SRT was observed with p-values ranging from p < 0.0001 to p = 0.0438.

## 4.0 Discussion

Our meta-analysis revealed that the LC in studies utilizing US-SRT was statistically superior to all four non-image-guided-XRT/SRT studies and this finding also held true when compared to all XRT/SRT studies combined. The four XRT/SRT articles were recent, high-quality with large sample sizes, and distilled from a previous ASTRO literature review. It was imperative that the studies we compared were comparing “apples to apples,” which is why only four studies utilizing XRT/SRT were included. Excluded comparator studies included advanced stage lesions that would have decreased overall LC and automatically skewed comparisons in favor of the US-SRT studies, which did not include advanced stage lesions (only T_is_, T_1_, & T_2_ lesions were studied).

This statistically significant improvement in LC with US-SRT can be attributed to the use of a dermal HRUS. HRUS allows the provider to visualize the skin tumor before, during, and after NMSC treatment. US visualization before treatment allows for the determination of tumor breadth and depth for width, energy, and dose selection. HRUS use during treatment allows the provider to make necessary adjustments in real-time, and after treatment allows for confirmation of treatment response. HRUS technology is easy to use, cheap, portable, and safe, as it does not use ionizing radiation. This technology offers patients a great benefit, especially when a patient is opting for non-surgical treatment of their NMSC lesion [18]. Confounding biologic dosing differences as a reason for the statistically significant improvement in LC with US-SRT instead of the image guidance factor was considered and determined to be unlikely since we calculate the biologic equivalent dose range in the four XRT/SRT papers’ dose regimens using TDF^A^ method and were determined to have similar or even higher biological doses than in the US-SRT studies.

In addition to the meta-analysis presented in this article, significance levels for similar comparisons were obtained using logistic models [19]. The statistical results and medical conclusions were substantively identical to those presented here, thereby validating the meta-analytical findings using an alternate statistical methodology. Our meta-analysis confirms and concurs with the log regression study, showing that US-SRT is statistically superior to the comparator XRT/SRT studies individually, collectively, and stratified by histologic subtype.

Recent studies show that MMS can provide 5-year local control rates of up to 99% for primary BCC and 92-99% for primary SCC [10, 20]. Further, recurrent BCC and SCC treated with MMS were shown to have control rates of 90-93% and 90%, respectively [10, 19]. For newly-diagnosed early-stage BCC and SCC, MMS was found to be superior for the treatment of SCC (local control rate of 98% vs. 75%, p < .01), but comparable for the treatment of BCC (98% vs. 96%, p = 0.26), however, only when compared to definitive radiotherapy [21]. Further, previous studies have examined the efficacy of RT on advanced NMSC, hence cure rates ranged from 50-100% [12, 22, 23]. It is important to compare “apples to apples” as there is potential for confounders to impact the interpretation of findings. Thus, a head-to-head randomized prospective match cohort study comparing US-SRT to MMS may be worthwhile to perform; although this may be difficult to accomplish as patients may be hesitant to be randomized to a surgical versus non-surgical arm.

The published US-SRT results separated by histology used in this study are comparable to MMS for the treatment of early-stage NMSC showing 99.1% and 98.9% control rates (Yu and Maloney, respectively) for BCC and control rates of 99.3% and 99.2% (Yu and Maloney, respectively) for SCC. This suggests that US-SRT is equivalent to MMS for the treatment of early-stage NMSC and potentially better long-term than MMS for SCC treatment.

## 5.0 Limitations

No randomized controlled trial exists for direct comparison of US-SRT to radiotherapy modalities, including superficial and external radiotherapy. The follow-up periods in this paper, though long enough to reasonably assure meaningful and accurate US-SRT to XRT/SRT comparisons, are unequal among studies.

## Data Availability

Deidentified data are available on request from the corresponding author: Dr. Lio Yu at. Data will be available for request with publication for period of at least 1 year.
Additional analyses may be available upon request, including but not limited to all odds and probabilities using logistic regression, conversion of some of the odds to probabilities with asymmetric confidence limits, raw percentages, and raw odds

## Statements and Declarations

### Ethics Approval

The study protocol was reviewed by an IRB committee (WIRB-Copernicus Group) and determined to be exempt from IRB approval under 45 CFR 46.104 (d)(4) as it fulfilled one or more of the exemption categories. Specifically, the information obtained is recorded by the investigator in such a manner that the identity of the human subject cannot readily be ascertained directly or through identifiers linked to the subjects, the investigator does not contact the subjects, and the investigator will not re-identify subjects. Any health information used in this study has been de-identified for use in this study. This study was performed in compliance with the pertinent sections of the Helsinki Declaration and its amendments. All methods were carried out in accordance with relevant guidelines and regulations.

### Author Contributions

Drafts of the manuscript were shared among the authors. All authors read and approved the final manuscript.

Dr. Lio Yu is responsible for conceptualization of the study design, methodology, data curation, funding acquisition, project administration, writing – original draft, writing – review & editing, supervision, investigation, visualization, and resources. Mairead Moloney was responsible for the literature search, writing – original draft, and writing – review & editing, and visualization. Dr. Alison Tran was responsible for writing original draft– review & editing. Songzhu Zheng was responsible for formal analysis, validation, and software. Dr. James Rogers was responsible for formal analysis, validation, software, writing – review & editing, figures, investigation, and visualization.

Songzhu Zheng directly accessed and verified the underlying data reported in the manuscript. Dr. James Rogers had access to the underlying data and applied statistical analysis to the data in tabulated format. The meta-analysis was validated under Summit Analytical SOPs.

### Role of Medical Writer or Editor

No medical writer or editor was used or involved in preparing this paper for publication.

## Data Availability

Deidentified data are available on request from the corresponding author – Dr. Lio Yu at. Data will be available for request with publication for period of at least 1 year.

Additional analyses may be available upon request, including but not limited to all odds and probabilities using logistic regression, conversion of some of the odds to probabilities with asymmetric confidence limits, raw percentages, and raw odds.

## Footnotes

A Time Dose Fractionation (TDF) is a useful system of tables representing biologic equivalent doses often used by radiation oncologists / therapeutic radiologists specifically for treatment of skin cancers with different fractionation schedules.

